# Implementation of pooled testing of sputum specimens from multiple individuals for rapid molecular detection of tuberculosis in Cameroon: retrospective evaluation of efficiency, cost, and instrument time to result

**DOI:** 10.64898/2026.03.20.26348676

**Authors:** Ngha Ndze Mbuh, Zourriyah Adamou Mana, Joceline Konso, Nankouo Arthur, Armand Toukap, Myriam Baiguerel, Neh Angela, Irene Adeline Goupeyou Wandji, Maurice Ganava, Ousmanou Bello, Mercy Fundoh, Meoto Paul, Fitame Adeline, Norah Nyah Ndi, Vuchas Comfort, Pride Teyim, Valerie Flore Donkeng Donfack, Tushar Garg, Jacob Creswell, Cyrille Mbuli, Melissa Sander, the INSPIRE TB team

## Abstract

**Background:** Only 54% of people with TB had an initial molecular diagnostic test in 2024, due to barriers including high test costs. Pooled testing is implemented in Cameroon as a strategy to increase testing efficiency on the Xpert MTB/RIF Ultra (Ultra) assay and extend molecular testing to more people when test reagents are limited, as recently recommended by the World Health Organization. At GeneXpert sites, laboratory personnel decide whether to test individually or in pools and pool size based on locally available information, including smear microscopy results, lab positivity rates, daily testing volume, availability of Ultra cartridges and GeneXpert modules, and patient characteristics.

**Methods:** We conducted a retrospective evaluation of Ultra testing at GeneXpert laboratories that implemented both individual and pooled testing in pools of 2 to 8. Ultra test results and duration were extracted from GeneXpert instruments. Testing efficiency, instrument time to result, and assay cost were analyzed overall and by pool size.

**Results:** From October 2023 to March 2025, 71,328 sputum specimens were tested at 16 GeneXpert laboratories. For 59,164 specimens tested in pools, including 1,999 (3.4%) with TB detected, 20,838 Ultra cartridges were used, or 0.35 cartridges per result, enabling an additional 38,326 people to have molecular test results compared to if specimens were tested individually. The average time to result varied from 45 minutes to 10 minutes for specimens tested in pools of 2 or 8, respectively, as compared to 66 minutes for individual testing. The calculated assay cost per result was $2.81 for specimens tested in pools (from $5.29 to $1.19 for specimens in pools of 2 or 8, respectively) as compared to $7.97 for individual testing.

**Discussion:** Implementation of pooled testing enabled many more people to have a molecular test result for TB, with significant time and cost savings compared to individual testing.

**What is already known on this topic:** Pooled testing is a strategy used to increase testing efficiency by combining specimens from multiple individuals prior to testing; if the pool tests negative, a negative result is reported for each specimen with no further testing, and if the pool tests positive, then each specimen from the pool is re-tested individually and the individual result is reported. The World Health Organization has recently recommended the use of pooled testing to increase access to molecular diagnostic testing for tuberculosis when resources are constrained.

**What this study adds:** This is the first report of large-scale programmatic implementation of pooled testing for the detection of TB. Pooled testing was performed by laboratory personnel who decided whether to test individually or in pools of size 2 to 8, based on the information available to them. Implementation of pooled testing enabled many more people to be tested with existing resources, with significant reductions in time to result and assay cost per specimen tested.

**How this study may affect research, practice or policy:** This demonstration of the successful scale up of pooled testing for TB should contribute to uptake of pooled testing by national TB programs and laboratories to increase access to molecular testing for TB when resources are constrained.

## INTRODUCTION

Many people with TB are never diagnosed and treated for the disease, with an estimated 2.4 million people with TB not linked to care in 2024.^1^ This gap in care is primarily responsible for the 1.25 million deaths from TB in 2024 and is in part attributable to lack of access to sensitive diagnostic tests for TB. The World Health Organization (WHO) reported that in 2024, only 54% of people treated for TB had a WHO-recommended, molecular test as the initial diagnostic test for TB.^1^ Many people in high TB burden settings are instead tested for TB using smear microscopy, which is inexpensive but has poor sensitivity to detect TB.^2^ Barriers to access sensitive molecular tests for TB include the high cost of the assays (typically $8 or more) and often low molecular testing capacity due to infrastructure and equipment limitations.^3,4^

Increased access to rapid molecular diagnostics is possible with pooled testing, an approach to increase testing efficiency by reducing the number of tests needed as compared to testing each person with a single test. In pooled testing as described by Dorfman, samples from multiple individuals are first pooled together and tested with a single assay; if the pool result is negative, all specimens are reported as negative, while if the pool result is positive, each specimen is then re-tested individually and the individual result is reported to the client.^5^ Pooled testing efficiency depends on the prevalence of the disease in the population tested, and it is more efficient than individual testing when the test positivity rate is relatively low, typically less than 25%.^5^ Pooled testing has been used for blood screening, disease surveillance, and detection of infectious diseases, including for COVID-19,^6–10^ HIV viral load,^11,12^, congenital cytomegalovirus (cCMV),^13,14^ and chlamydia and gonorrhea.^15^

Pooled testing for TB using the Xpert MTB/RIF and Xpert MTB/RIF Ultra assays (Ultra, Cepheid, USA) has shown promising improvements in efficiency in multiple research studies.^16–24^ Supported by evidence from a large multi-country diagnostic accuracy study against a TB culture reference standard, pooled testing for TB was recently recommended as a testing strategy by WHO for pools of up to 4 specimens.^25^ Pooled testing on the Ultra cartridge has been demonstrated to detect ≥90% of specimens with TB as compared to individual testing.^26^ The Ultra assay is based on detection of multi-copy targets (IS1081 and IS6110) in the *Mycobacterium tuberculosis* complex DNA as compared to the earlier Xpert assay that was based on detection of a single-copy target (rpoB gene).^27^ For pooled testing on the Ultra assay, the observed increase in the cycle threshold as compared to individual testing is lower than the theoretical (log_2_(n), where n is the pool size), leading to better agreement with individual test results for this multi-copy target assay on pooled testing as compared to the single-copy target Xpert assay.^16,26,28^

While implementation of pooled testing has been described for other diseases, there are not yet reports of large scale, programmatic implementation of pooled testing for TB. In Cameroon, pooled testing for TB was initially adopted at reference laboratories in 2020 to address shortages in test cartridges during the COVID-19 pandemic^28^, and it was subsequently scaled up to additional GeneXpert laboratories, building on a previous TB case finding intervention in six regions of the country.^29^

In this work, we report a retrospective evaluation of pooled testing implemented at a network of GeneXpert labs as part of an intervention to improve TB detection. We describe the process used to implement pooled testing for the detection of TB with pools of 2 to 8 specimens on the Ultra assay, and we report pooled testing efficiency, instrument time to results and test cost per specimen.

## METHODS

We conducted a retrospective evaluation of laboratory data that was collected during Ultra individual and pooled testing from October 2023 to March 2025. Pooled testing was implemented to increase TB case detection as part of a TB REACH intervention. Testing was conducted at GeneXpert laboratories using the Ultra assay across 6 of the 10 regions in Cameroon, including the Far North, North, West, Littoral, Southwest and Northwest regions (Figure 1); labs that tested >100 specimens in pools during the period were included in the analysis. Sputum specimens were collected at 183 facilities that referred specimens from people with presumptive TB for TB testing.

**Figure 1.**
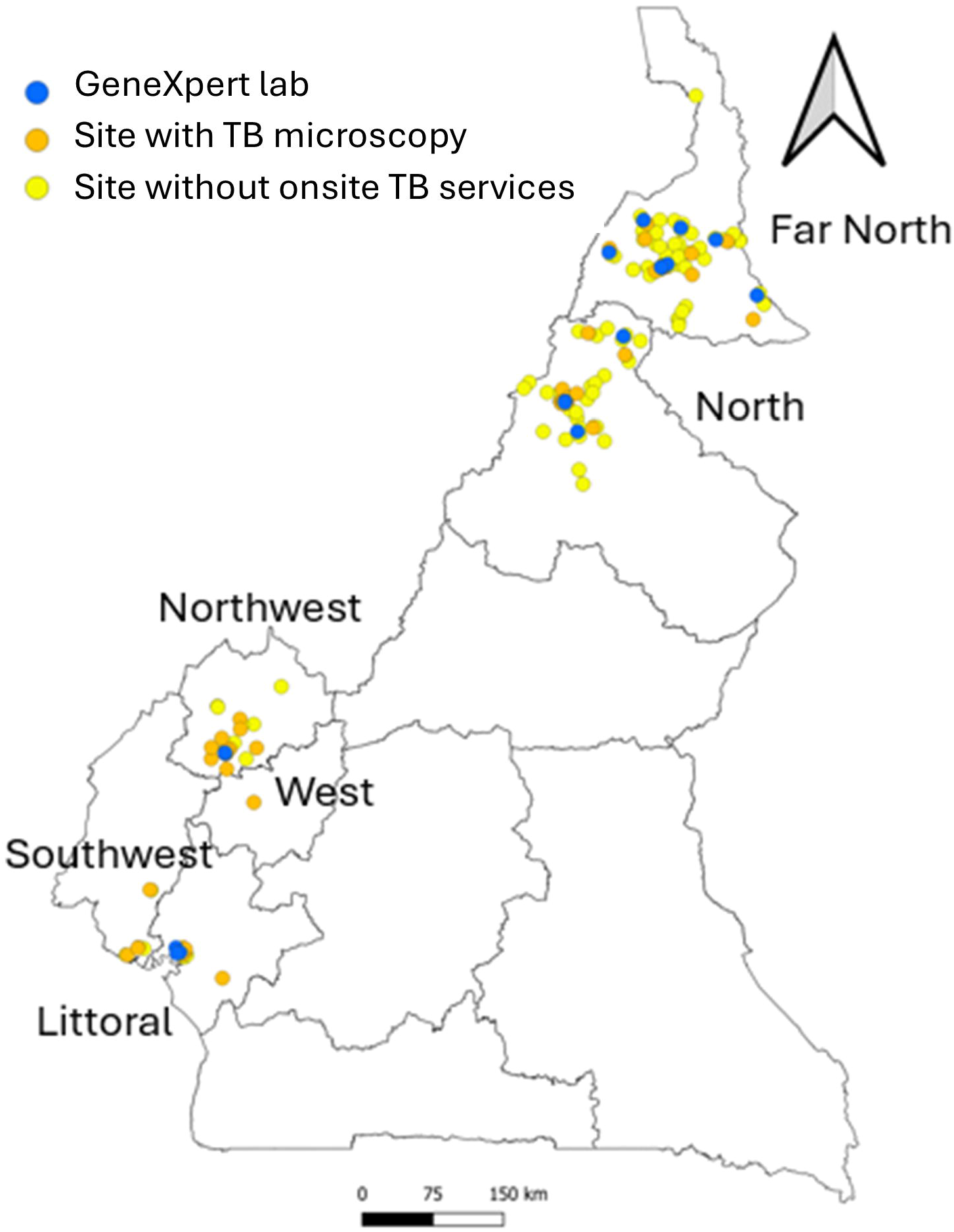
Map of Cameroon, showing the 183 health facilities, including 16 with GeneXpert labs (blue), 53 sites with microscopy for TB testing (orange) and 114 without TB diagnostics (yellow), that referred specimens for Ultra testing, across 6 of the 10 regions of the country.

Depending on the laboratory techniques conducted onsite or at the referral testing facilities, sputum specimens were tested with TB microscopy, TB LAMP (Eiken, Japan) and/or Ultra following national algorithms for TB testing and using existing specimen referral networks as well as specimen referral pathways developed as part of the intervention. Most specimens were collected at sites without molecular testing capacity, so smear microscopy was often performed first at the collection site or at the nearest TB diagnostic and treatment center with microscopy services, prior to receipt at the molecular testing laboratory. If smear microscopy was negative but the molecular test later confirmed TB, the client was contacted to initiate treatment based on the molecular result. In line with national guidelines, typically only one sputum specimen per person was tested using a rapid diagnostic method (Ultra or TB LAMP).

The intervention was approved by and conducted in collaboration with the Cameroon National TB Program. All testing data was de-identified prior to analysis. All specimen testing methods were carried out in accordance with approved guidelines. The study is reported following the REporting of studies Conducted using Observational Routinely-collected health Data (RECORD) guidance^30^.

### Pooled testing implementation

Pooled testing was implemented on a pragmatic basis. Laboratory technicians with prior experience performing individual Ultra testing for TB were trained on pooled testing in a one-day workshop facilitated by reference laboratory and National TB Program personnel. Indicators on pooled testing performance including efficiency were monitored continuously and reviewed in monthly virtual meetings, and implementers also participated in a WhatsApp group to share information. Refresher training and training of new personnel was conducted as needed for individual technicians and laboratories.

Laboratory personnel at each GeneXpert site determined whether to test specimens individually or in pools, as well as the appropriate pool size. These decisions were guided by several factors: the availability of smear microscopy results (specimens confirmed smear-positive were tested individually and smear-negative specimens were often pooled), operational considerations such as historical positivity rates, daily testing volume, turnaround time requirements, cartridge availability, and instrument functionality. Specimens less than 1mL in volume were typically tested individually. When available, patient-level information—such as symptom profile, treatment history, and HIV status—also informed pooling decisions. (Figure 2a)

**Figure 2.**
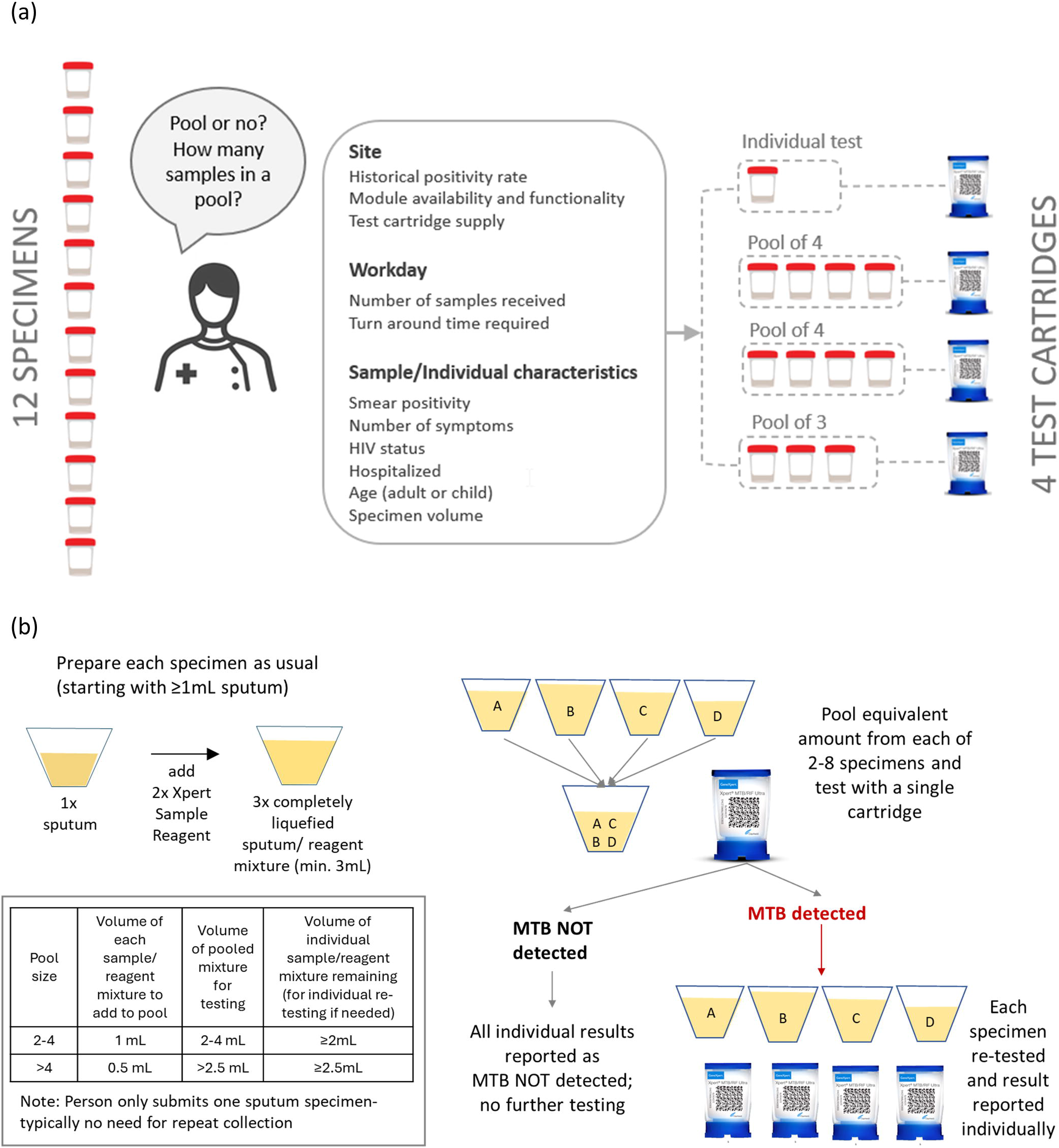
Implementation of pooled testing for the detection of TB. a) Laboratory personnel decide on whether to pool or not and how many specimens to add to the pool depending on local information available to them on a daily basis. In this illustration, at a lab that typically tests in pools of 4 based on historical positivity rates, they may test one specimen individually (for example, a specimen from a child or a smear positive specimen), 2 pools of 4 and then 1 pool of 3, if they receive 12 specimens in a day. (b) Schematic of pooled testing as conducted on the Xpert MTB/RIF Ultra (Ultra) assay.

### Sputum specimen processing and creation of pools for testing

The detailed procedure is summarized in a 5-minute video (available at: https://youtu.be/gQdQ60PbAhQ) and job aid (Supplemental Information) and has been described previously.^28^ Pooling was typically only conducted on sputum specimens that had at least 1mL of sputum; this is also the minimum volume recommended by the manufacturer for the test.^27^ Sputum specimens were initially processed following the manufacturer’s recommendation; briefly the Xpert Sample Reagent was added in a ratio of 2:1 to the sputum in the sputum collection container, the mixture was closed and shaken vigorously then allowed at room temperature for 15 minutes, or for additional time if needed until fully liquefied.

To create pools for testing, a new sputum container was used for the pool, and an equivalent amount of each specimen-reagent mixture was pipetted into the new container to give a minimum of 2mL for the pool (Figure 2b).The leftover specimen-reagent mixture for each sample (with a minimum volume of 2mL) was set aside and used for individual testing in case of a pool result that was positive or indeterminate. The specimen-reagent mixture was stored and re-tested following the manufacturer’s guidance of within 4 hours if stored at room temperature, or within 24 hours if stored in the refrigerator at 2–8 °C.

To perform pooling, one additional sputum mug was required for each pool (to create the pool) and one disposable pipet was required for each specimen to be pooled. These additional consumables were procured locally, at 100FCFA ($0.18) per sputum mug and 80FCFA ($0.14) per disposable pipet.

### Ultra testing for pooled and individual specimens

To perform the assay, slightly more than 2mL of the specimen-reagent mixture was added to a single Ultra cartridge, and the specimen was run on the GeneXpert (GX) instrument following the standard procedures for testing. For specimen pools with a result of MTB NOT DETECTED, all individual specimen results from the pool were then reported as MTB NOT DETECTED. For specimen pools with any result of MTB DETECTED, each specimen was re-run individually and that individual result was reported as the final individual specimen result. For pools with indeterminate results, either the individual samples or the pool was subsequently re-tested, as decided by the lab personnel based on the error code and the amount of remaining specimen-reagent mixture for the pool. In case of a positive pool followed by all negative results on individual tests, new specimens from individuals in the pool were requested and tested, with additional follow-up as needed.

### Data extraction and preparation

People eligible for a TB test were typically assigned a unique Quick Response (QR) code that was scanned into the REDCap Mobile App^31^ during screening and affixed to the sputum mug for specimen collection. At the GeneXpert lab, this QR code, or if the QR code was not available, another lab-assigned unique number for the specimen, was entered into the GX instrument; this was done by scanning the QR code with the GX scanner or by manually typing the number during specimen accessioning prior to testing. For pools, the QR code or other unique specimen number from every specimen in the pool were entered together, typically in the ‘Notes’ field for the Ultra test result. Pooled tests with positive results were matched to subsequent respective individual test results using the unique QR code or other unique lab number.

Ultra test results were extracted from the GeneXpert instrument. Tests with a result of MTB DETECTED (any grade) were classified as positive, tests with a result of MTB NOT DETECTED were classified as negative, and tests with a result of error, invalid or no result on the assay were classified as indeterminate. Only tests with positive or negative results and with unique IDs that were successfully linked between pool and individual results were retained for analysis; the number of pooled and individual tests with indeterminate results were summarized separately.

The duration for each test (instrument time to detection) was obtained from the GeneXpert instrument (e.g. end time minus start time). The Ultra assay time to result ranges from approximately 65 to 87 minutes, with results of MTB not detected or MTB detected Trace closer to 65 minutes and results with MTB detected very low to high taking longer due to the detection of rifampin resistance on these specimens^32^.

Lab data including test results and time to result for each specimen tested on Ultra was compiled using the unique serial numbers of the Ultra cartridges, together with the unique QR code or other unique lab-assigned identification number and the pool number, when pooled, to link the specimen result to the pool result.

### Statistical analysis

The number of overall pools and specimens tested, the number of pools and specimens that tested positive, and the total number of Ultra tests performed were summarized, both overall and stratified by pool size.

The percentage of cartridges saved due to pooled testing as compared to individual testing was estimated by dividing the number of tests used for pooled testing by the number of cartridges that would have been needed to test all specimens individually.

Statistics on GeneXpert lab testing, including numbers of TB specimens tested per day and lab positivity rates, were summarized from the extracted Ultra testing data from the GeneXpert instrument.

Time to result was calculated as instrument time taken to obtain the final test result for each specimen; for specimens tested in pools, this included the time for pooled and follow-on individual tests for specimens in positive pools and time for pooled tests for specimens in negative pools. The time to result described here is the instrument time and does not include the time to prepare the specimen for testing^33^. For the small proportion of results (<0.3%) that were out of the range of 63-74 minutes for MTB not detected or MTB trace results, or out of the range 74-90 minutes for MTB detected very low or above, the outlier values were replaced with the median time for result for that category.

To evaluate the agreement between the cycle threshold (Ct) values for specimens that tested positive initially in a specimen pool and subsequently by individual testing, we used the Bland-Altman method^34,35^. Only specimens in pools with a single positive result were included in this analysis.

All analyses were performed in R version 4.5.0; the method comparison analyses were performed using the ‘mcr’ package ^36^. Other calculations are shown in Table 1.

**Table 1.** Testing statistics, efficiency, time to result, and assay cost of pooled testing of 59,164 specimens with the Xpert MTB/RIF Ultra (Ultra) assay, by pools of size 2 to 8, and overall.

| Pool Size | 2 | 3 | 4 | 5 | 6 | 7 | 8 | All |
| --- | --- | --- | --- | --- | --- | --- | --- | --- |
| **Summary statistics** |  |  |  |  |  |  |  |  |
| Number of pools with results | 1,378 | 2,401 | 6,184 | 1,152 | 2,546 | 167 | 283 | 14,111 |
| Number of pools with MTB detected | 226 | 369 | 827 | 102 | 204 | 10 | 7 | 1,745 |
| Number of specimens with test results | 2,756 | 7,203 | 24,736 | 5,760 | 15,276 | 1,169 | 2,264 | 59,164 |
| Number of individual tests (after positive pool) | 452 | 1,107 | 3,308 | 510 | 1,224 | 70 | 56 | 6,727 |
| Number of positive specimens | 223 | 423 | 981 | 119 | 236 | 10 | 7 | 1,999 |
| Positivity rate (% specimens with MTB detected) | 8.1% | 5.9% | 4.0% | 2.1% | 1.5% | 0.9% | 0.3% | 3.4% |
| Number of PCR tests (cartridges) used | 1,830 | 3,508 | 9,492 | 1,662 | 3,770 | 237 | 339 | 20,838 |
| **Efficiency of pooled testing** |  |  |  |  |  |  |  |  |
| Number of tests run per person with result (tests / specimens with results) | 0.66 | 0.49 | 0.38 | 0.29 | 0.25 | 0.20 | 0.15 | 0.35 |
| Estimated additional people with test results (vs. individual testing) | 926 | 3,695 | 15,244 | 4,098 | 11,506 | 932 | 1,925 | 38,326 |
| Estimated % of assays saved with pooled testing (vs. individual testing) | 34% | 51% | 62% | 71% | 75% | 80% | 85% | 65% |
| **Time to result- instrument (minutes)** |  |  |  |  |  |  |  |  |
| Measured instrument time to result- pooled testing (minutes) |  |  |  |  |  |  |  |  |
| Total instrument time- all tests (20,838 tests, n= 59,164) | 125,214 | 238,260 | 643,625 | 111,263 | 251,228 | 15,749 | 22,581 | 1,407,920 |
| Average time per specimen in negative pool (n= 52,437) | 33.0 | 22.0 | 16.5 | 13.1 | 10.9 | 9.4 | 8.3 | 15.5 |
| Average total time pool + individual per specimen in positive pool (n= 6,727) | 108.7 | 94.3 | 87.8 | 83.0 | 79.9 | 77.6 | 77.1 | 88.3 |
| Average instrument time per specimen in pools (n = 59,164) | 45.4 | 33.1 | 26.0 | 19.3 | 16.4 | 13.5 | 10.0 | 23.8 |
| Estimated instrument time to result- individual testing (minutes)* |  |  |  |  |  |  |  |  |
| Total estimated instrument time- all tests (n= 59,164) | 183,674 | 478,507 | 1,638,708 | 380,534 | 1,008,441 | 77,094 | 149,188 | 3,916,145 |
| Average estimated instrument time per individual specimen (n = 59,164) | 66.6 | 66.4 | 66.2 | 66.1 | 66.0 | 65.9 | 65.9 | 66.2 |
| Estimated % instrument time saved with pooled testing (vs. individual testing) | 32% | 50% | 61% | 71% | 75% | 80% | 85% | 64% |
| **Assay cost (USD)** |  |  |  |  |  |  |  |  |
| Total cost- cartridges used for pooled testing (n= 20,838 \$7.97/cartridge) | \$14,585 | \$27,959 | \$75,651 | \$13,246 | \$30,047 | \$1,889 | \$2,702 | \$166,079 |
| Assay cost per specimen with result- pooled testing | \$5.29 | \$3.88 | \$3.06 | \$2.30 | \$1.97 | \$1.62 | \$1.19 | \$2.81 |
| Estimated cost- cartridges for individual testing (n= 59,164 \$7.97/cartridge) | \$21,965 | \$57,408 | \$197,146 | \$45,907 | \$121,750 | \$9,317 | \$18,044 | \$471,537 |
| Assay cost per specimen with result- individual testing | \$7.97 | \$7.97 | \$7.97 | \$7.97 | \$7.97 | \$7.97 | \$7.97 | \$7.97 |
| Estimated % cartridge cost saved with pooled testing (vs. individual testing) | 34% | 51% | 62% | 71% | 75% | 80% | 85% | 65% |

\* Using average measured time per negative test of 65.9 minutes and average measured time per positive test of 75.5 minutes (Figure S1)

## RESULTS

Specimens tested at 16 GeneXpert labs from October 1, 2023 to March 31, 2025 were included in this analysis, including those tested by individual or pooled testing (Figure 3). In total, 71,328 specimens were tested either individually (12,164 specimens, 17%) or in 14,111 pools (59,164 specimens, 83%) with valid and complete results. Of samples tested individually, 661 (5.2% of 12,825) had initial indeterminate results (including 489 with results of error, 60 invalid, and 112 no result), and 503 pools tested (3.3% of 15,260) had initial indeterminate results (with 415 error, 20 invalid, 68 no result); the samples with indeterminate results were subsequently re-tested in pools or individually. Among 14,757 pools with valid results, 418 positive pools had 1 or more incomplete results as the pool result was unable to be matched to all subsequent individual results, and 228 pools had one or more indeterminate result. Only tests with valid and complete results were included in the analyses.

**Figure 3.**
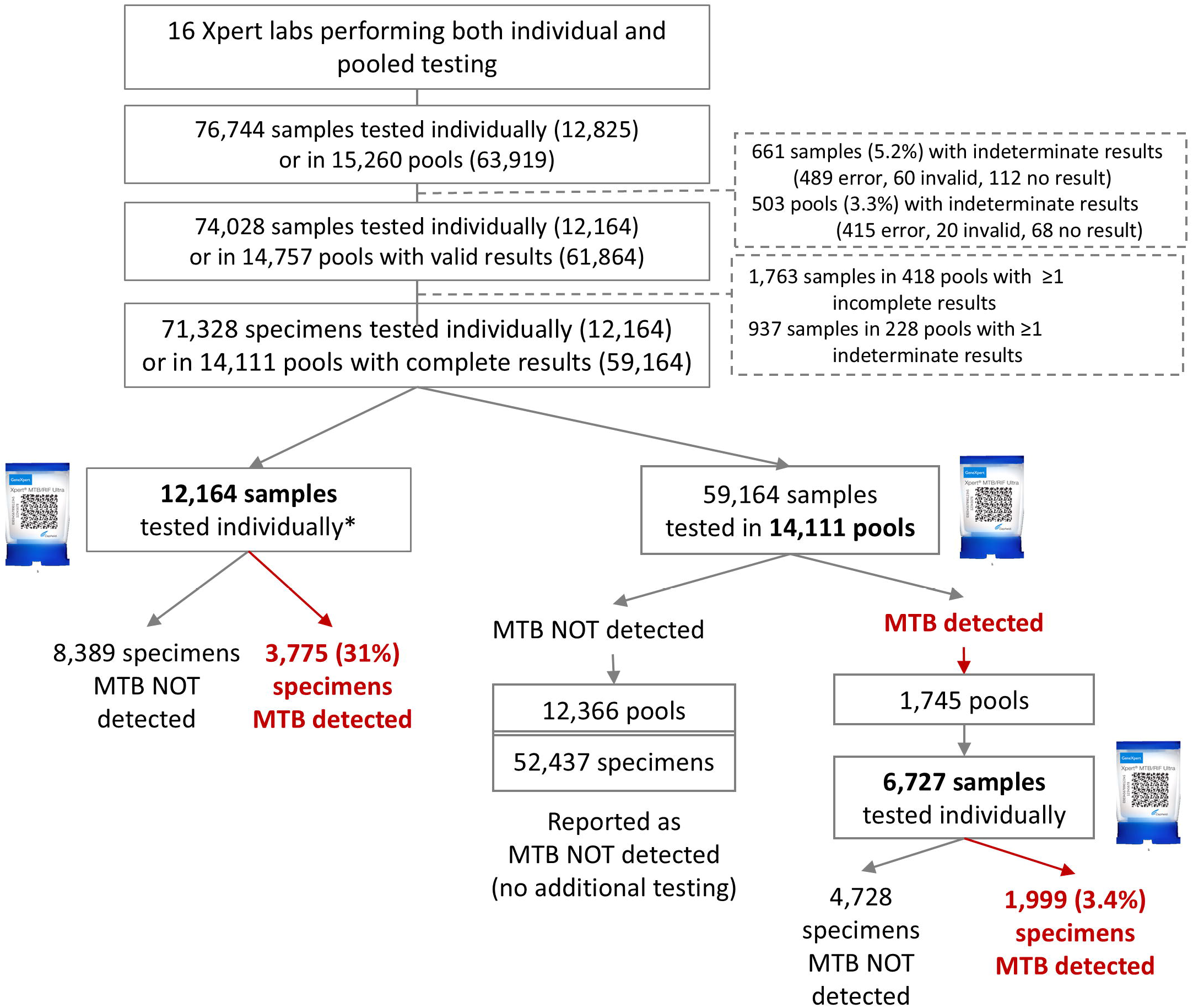
Flow of individual and pooled testing on Xpert MTB/RIF Ultra (Ultra) assay. *Many specimens had smear microscopy results at the initial testing lab prior to referral to the GeneXpert testing lab. Smear positive specimens were typically tested on Ultra individually. MTB, *Mycobacterium tuberculosis* complex

### Pooled testing: Efficiency

The efficiency of testing in this population was assessed overall and by pool size, as shown in Table 1. Overall, 59,164 specimens were tested using 20,838 cartridges, or 0.35 cartridges per result obtained, saving 65% of cartridges. These specimens were tested in 14,111 pools; of these, 1,745 pools (12.4%) had MTB detected with 1,999 individual specimens (3.4% of pooled specimens tested) that had MTB detected. Most positive pools had a single positive specimen (73%, 1,271/1,745), while 293 pools (17%) contained 2 positive specimens, 35 pools (2%) contained 3 positive specimens, 8 pools (0.5%) contained 4 positive specimens. Overall, 137 pools (7.8% of positive pools) had a positive result with none of the specimens tested individually testing positive; of these, 86 pools (62%) had a result of MTB Trace, and 41 (30%) had results of MTB low or very low.

Testing efficiency varied by pool size and specimen positivity. For pools of 2, with 8.1% of specimens positive, 0.66 cartridges were used per result obtained, while for pools of 4 with 4.0% of specimens positive, 0.38 cartridges were used per result obtained. For pools of 5 to 8, with 2.1% to 0.3% of specimens positive, efficiency was from 0.29 to 0.15 cartridges used per result.

### Pooled testing: Instrument time to result

From the instrument time reported by the instrument for the 59,164 specimens tested in 14,111 pools and 6,727 subsequent individual tests, the average instrument time per specimen was 23.8 minutes; for specimens in negative pools, the average instrument time per result was 15.5 minutes and for specimens in positive pools with a subsequent individual test, the average instrument time was 88.3 minutes. The average time to result decreased as pool size increased; pools of 2 averaged 45.4 minutes per specimen with result compared to pools of 8 (10.0 minutes).

Using pooled testing, a total of 41,804 hours (2,508,224 minutes) of instrument time was saved as compared to individual testing. The overall time taken was 64% less than what would have been necessary if all specimens had been tested individually. For specimens tested individually after a positive pool result, the median instrument time for 5,063 results of MTB not detected or MTB detected trace was 65.6 minutes (range, 63.7-71.8 minutes), while the median instrument time for 1,664 results with MTB detected grades of very low to high was 77.4 minutes (range, 75.3-84.5 minutes). The time to positive and negative result among specimens with a positive individual result following a positive pool was similar to the 65.9 minute and 77.5 minute median times for negative and positive specimens, respectively, tested individually (Figure S1 and Table S2).

### Pooled testing: Assay cost

For the 59,164 specimens tested in pools, a total of $166,079 was spent on the 20,838 cartridges with valid results ($7.97 /cartridge^37^); if these specimens had been tested individually, the total cartridge cost would have been $471,537 (59,164 cartridges at $7.97/cartridge). The cost per specimen with result averaged $5.29 for specimens tested in pools of 2, $3.06 for specimens tested in pools of 4, $1.96 for specimens tested in pools of 6, and $2.81 overall.

### Individual or pooled test and pool size, by GeneXpert lab characteristics

Specimens tested in labs with lower positivity rates and higher numbers of specimens tested per day were more likely to be tested in pools (Table 2). In laboratories that tested <10 specimens per day, 65% of specimens (8,473/13,046) were tested in pools, as compared with 90% of specimens (39,088/43,267) tested in pools in labs that tested >20 specimens per day. In labs with a test positivity rate of 12-15%, 33% (6,578/20,174) of specimens were tested individually as compared to 8% (1,232/15,466) of specimens tested individually in labs with a positivity rate of 3-5%.

**Table 2.** Specimens tested individually or in pools on the Xpert MTB/RIF Ultra assay.

|  | Individual |  | Pooled |  | Total |
| --- | --- | --- | --- | --- | --- |
|  | n (%) |  | n (%) |  | n |
|  | 12,164 | (17) | 59,164 | (83) | 71,328 |
| **Laboratory characteristics** |  |  |  |  |  |
| Number of specimens tested per day |  |  |  |  |  |
| <10 | 4,573 | (35) | 8,473 | (65) | 13,046 |
| 11-20 | 3,412 | (23) | 11,603 | (77) | 15,015 |
| >20 | 4,179 | (10) | 39,088 | (90) | 43,267 |
| Overall test positivity rate |  |  |  |  |  |
| 3-5% | 1,232 | (8) | 14,234 | (92) | 15,466 |
| 6-8% | 1,844 | (9) | 19,344 | (91) | 21,188 |
| 9-11% | 2,510 | (17) | 11,990 | (83) | 14,500 |
| 12-15% | 6,578 | (33) | 13,596 | (67) | 20,174 |

Among specimens tested in pools, a total of 42% (24,736/59,164) were tested in pools of 4 and 26% (15,276/59,164) were tested in pools of 6 (Table S1). Larger pool sizes were used in labs with lower overall positivity rates (47% of specimens tested in labs with 3-5% positivity were tested in pools of 6 as compared to 8% of specimens in labs with 12-15% positivity). The positivity rate among pooled specimens was 3.4% (1,999/59,164).

### Ultra results on pooled and individual testing

In total, 35% (2,044/5,774) of specimens with MTB detected had a grade of HIGH on the Ultra assay, including 43% (1,626/3,775) of positive specimens tested individually and 20% (408/1,999) of positive specimens tested individually after a positive pool result; among specimens with MTB detected following pooled testing, 65% (1,293/1,999) had Ultra grade of low, very low or trace, as compared to 38% (1,446/3,775) of positive specimens with these lower grades tested individually (Table S2).

From the Bland-Altman agreement analysis, the mean bias between the cycle threshold of the pooled and individual results varied from +0.20 cycles (95% limits of agreement, +4.38 to −3.97 cycles) for pools of 2 to 1.37 cycles (95% limits of agreement, +5.55 to −2.82 cycles) for pools of 6 (Figure S2). The standard deviation of the bias across all pools of 2 to 6 was 2.0 cycles.

## DISCUSSION

In this retrospective analysis of Ultra testing performed at 16 GeneXpert labs over 1.5 years, more than 80% of specimens were tested for TB in pools of 2 to 8 rather than individually. Pooled testing enabled an additional 38,326 people to access molecular test results for TB, using just 0.35 tests per result, and greatly reduced GeneXpert instrument time to results as compared to individual testing. These results may provide a useful guide for TB programs and laboratories to implement pooled testing for TB, following the WHO recommendation of pooled testing as a novel testing strategy to increase access to molecular testing.^25^

In this implementation of pooled testing on Ultra, the average assay cost per specimen with result was $2.81, which was 65% less than the $7.97 for an individual Ultra test, ranging from $5.29 for specimens tested in pools of 2 to $1.19 for specimens tested in pools of 8. New near point of care diagnostic tests for TB, such as the PlusLife MiniDock MTB, have been demonstrated to have sensitivities meeting the WHO target product profiles for peripheral labs^38,39^, and these tests are expected to be available at much lower cost than the $7.97 Ultra assay, with the current price of the PlusLife MiniDock MTB at $3.60 per test.^37^ Both pooled testing for TB and near point of care testing for TB can be used to increase access to molecular testing for TB by reducing the cost of testing. Since many countries already have GeneXpert instruments (and/or other low complexity nucleic acid amplification testing platforms) installed and testing for TB at scale, use of pooled testing is an approach that could have a rapid impact to extend access to molecular testing for many more people, using existing infrastructure and resources; this could be done in parallel with the roll-out of new near point of care TB tests as countries adopt these. Because external funding for global health is under immense pressure and budgets for TB diagnostics are being further reduced in many settings, the implementation of an approach that does not require additional investment may help mitigate insufficient funding for TB diagnostics.

Importantly, implementation of pooled testing as described here was adapted to local needs, with the decision to test individually or in pools, and size of pools, taken by lab personnel at the testing laboratories based on the information available to them. These results show that pooling is feasible in large scale programmatic settings across multiple labs at different levels of care, and decisions around workflow and management of samples can be routinely managed by laboratory personnel.^40^ In this setting, many specimens arriving at the GeneXpert labs had a previous test for microscopy at the local lab. To increase the efficiency of pooled testing, specimens with known positive microscopy results were tested individually on the Ultra assay.^41^ Other considerations to inform the decision to pool and pool size were historical laboratory positivity rates and number of specimens received each day or during the test result turnaround window for the client (Table 2); laboratories were empowered to adjust pool sizes based on specimen testing volume (eg. a pool of 4 and a pool of 5 if 9 specimens were received during the testing period (Figure 2A, Table S1). In general, labs used pool sizes that were smaller than the optimal pool size based on the lab positivity for specimens tested in pools (Table S1)^5^, and this practical, non–optimized implementation still led to significant increases in testing efficiency and many more people with results. During the implementation period, there were also frequent Ultra cartridge shortages and GeneXpert module failures that contributed to testing decisions. In addition, patient characteristics, including TB treatment history and TB symptoms, were often but not always available to the lab personnel, either on the testing request form provided by the National TB Program and/or in the REDCap Mobile App implemented as part of the intervention. Testing efficiency could be further improved by providing customized guidance to lab personnel, for example through an app that uses Informative pooled testing, as has been described previously, to guide the decision to pool or test individually and to optimize pool size^41–43^.

While the WHO has recommended pooled testing for low complexity nucleic acid amplification tests (LC-NAAT) such as the Ultra for pools of size 2 to 4 based on the data that was available at the time of the review,^25^ our results suggest that larger pools sizes are feasible and may be useful in some situations. The exploratory sensitivity analyses by Teles et al for pools of 8 on the Ultra assay indicated a positive percent agreement of >90% for pools of 8, although with a small sample size.^26^ In this work, approximately 36% of the 59,164 specimens tested in pools in this evaluation were tested in pools of 5 or 6, with an additional 6% tested in pools of 7 or 8. These larger pool sizes were performed mostly at laboratories with overall positivity rates of 3-8% (Table S1), and the test positivity rates among specimens pooled was 3.4% overall, corresponding to a theoretical optimal pool size of 6.^5,44^ Bland-Altman agreement analysis indicated greater dilution at the higher pool size as expected, but the positive percent agreement as compared to individual testing for pools of up to at least 6 is still expected to be high, since few specimens have TB detected at high cycle threshold values on the Ultra assay (Figure S2). Larger pools have the potential to be highly cost-effective and have good diagnostic value for high volume testing in resource constrained situations; further studies are needed to assess both diagnostic performance and cost effectiveness across a range of pool sizes and settings.

In addition to being able to test many more people, pooled testing saved 65% of GeneXpert instrument time as compared to individual testing. Instrument time savings can help to free up time for more testing on these instruments that are used for multi-disease testing, including for HIV viral load and early infant diagnosis of HIV testing. The faster test turnaround time also contributes to reducing the time to results for patients.

We found that almost 8% of pools with positive results had discordant individual results, with none of the specimens tested individually having a positive result. Similar rates of discordance (or ‘false’ positives) have been reported with other RT-PCR assays, including 14.5% in pooled testing for cCMV^13^ and 2.5-5.3% in pooled testing for SARS-CoV-2.^45^ These discordant results may be in part due to the variability of the assay; among 1,133 pools with a single positive in the pool, the standard deviation of the bias was 2.0 cycles (Figure S2). This is similar to the standard deviation reported by the manufacturer for the assay (1.7 cycles)^27^. Several factors can contribute to assay variation, including chemical efficiencies (of the enzymes, primer and/or template) and variable equipment factors including fluorescence detection, temperature and variations in reagent volume. Assay variability can lead to pool results with MTB detected followed by no positives in the individual tests from that pool, and it can also contribute to false negative pools. The clinical impact of discordant pools can be mitigated by follow-up of the individuals in the positive pool for repeat testing and clinical follow-up to ensure each person with a specimen in the pool either gets better or is re-evaluated for TB. Assay variability also contributes to missed detection of TB during individual testing.

This evaluation had several limitations. This was a retrospective evaluation of laboratory data, so other information relevant to decisions about pooling such as smear result, individual patient level data, reagent availability and equipment status were not included. In addition, information about people with specimens tested in discordant pools was followed up by the local labs but not documented systematically, so we are unable to report how many of these pools included a person who was eventually diagnosed with TB. Specimens were matched based on a unique ID, mostly a QR code but sometimes a lab unique ID, and errors in data entry led to exclusion of some positive pools in which the pooled and individual results could not be fully matched (Figure 3); use of unique ID for linking pooled to subsequent individual testing results for a specimen is critical for high quality data and in laboratory information systems. As a programmatic evaluation, we did not test each sample individually and cannot quantify how many people pooling may have missed by individual testing; instead, pooled testing was implemented because there were not enough Ultra cartridges available to provide individual tests to everyone who could benefit from them.

## Conclusion

Implementation of pooled testing to detect TB on the Ultra assay in six of the 10 regions of Cameroon was efficient, leading to significant cost and time savings and providing access to molecular testing for tens of thousands of people with presumptive TB who would not have received a molecular test result without pooled testing. Laboratory personnel who were trained to conduct pooled testing made independent decisions to improve efficiency, deciding whether to test individually or in pools and pool size based on local information and needs. Pooled testing can be implemented with existing resources and has the potential to be widely scaled up without additional investment. Use of pooled testing as a testing strategy when resources are constrained can help to ensure that many more people have access to sensitive molecular testing for TB, and potentially as well as for other diseases as testing becomes more integrated.

## Supporting information

Job Aid

## Data Availability

All data produced in the present study are available upon reasonable request to the authors

