## Supplementary material for "Implementation of pooled testing of sputum specimens from multiple individuals for rapid molecular detection of tuberculosis in Cameroon: retrospective evaluation of efficiency, cost, and instrument time to result": Figures and Table

Figure S2. Bland-Altman plots to compare cycle threshold values of specimens with positive results tested both in pools of size 2 and 6 and individually on the Xpert MTB/RIF Ultra assay .. 5

Table S3. Summary of Bland-Altman agreement by pool size and overall ................................... 5

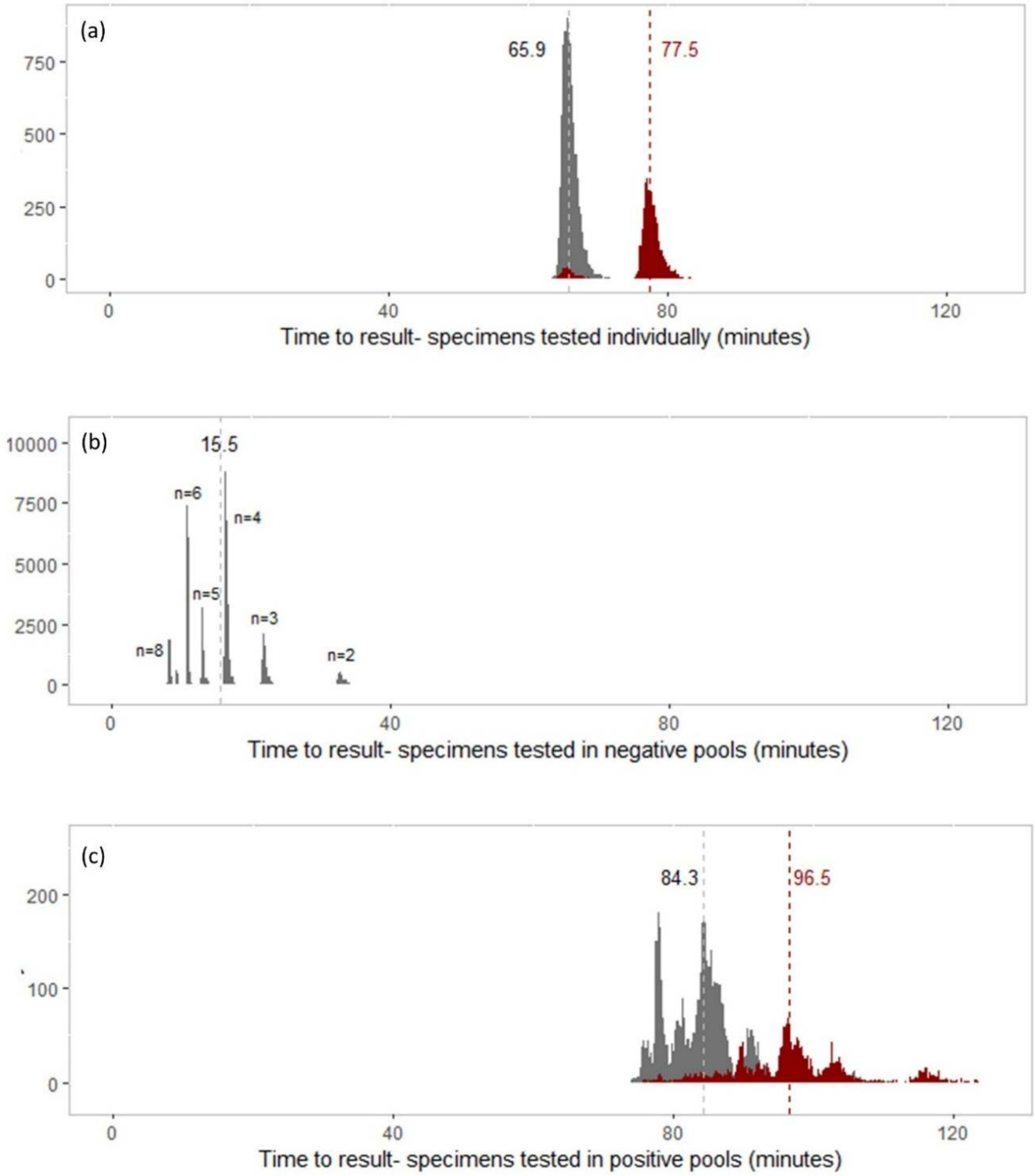

**Figure S1. Histograms of instrument time to test result in minutes.**

(a) specimens tested individually, (b) specimens tested in pools with negative results and (c) specimens tested in pools with positive results, including the time for the initial pool result plus the subsequent individual test result. Final results for specimens are shown as MTB Detected (red) and MTB not detected (grey). The dashed vertical lines indicate the average time for the respective subgroups. In panel B, the pool size (n) is shown for pools of size 2 to 8. Tests with time to results that were outside of the acceptable range were replaced with the median times; this included for (a) 7 results (7/12,164 = 0.1%) , for (b) 68 results (68/52,446 = 0.1%) and for (c) 19 (19/6,727 = 0.3%).

**Table S1. Proportion of specimens tested at each pool size on Xpert MTB/RIF Ultra assay**

| Lab overall positivity rate | Pool size 2 | 3 | 4 | 5 | 6 | 7 | 8 | Number tested in pools | Positivity rate, pooled specimens |
|---|---|---|---|---|---|---|---|---|---|
| 3-5% | 2% | 4% | 45% | 2% | 47% | | | 14,234 | 2.1% |
| 6-8% | 3% | 11% | 27% | 15% | 30% | 4% | 11% | 19,344 | 3.4% |
| 9-11% | 3% | 7% | 65% | 10% | 13% | 1% | 1% | 11,990 | 4.4% |
| 12-15% | 12% | 28% | 39% | 11% | 8% | 2% | | 13,596 | 3.8% |
| All | 5% | 12% | 42% | 10% | 26% | 2% | 4% | 59,164 | 3.4% |

The percentages in the body of the table indicate proportion of the total number of specimens tested in pools, by pool size, for each row of lab positivity category. Each row adds to 100%. The overall positivity rate includes all specimens tested, including both individual and pooled specimens.

**Table S2. Xpert MTB/RIF Ultra assay results, by grade, for specimens tested individually and in pools**

| | Total | | Tested individually | | Tested in pools | |
|---|---|---|---|---|---|---|
| | n (%) | | n (%) | | n (%) | |
| | 71,328 | | 12,164 | (17) | 59,164 | (83) |
| **Ultra results with grade** | | | | | | |
| MTB Detected | 5,774 | (8) | 3,775 | (31) | 1,999 | (3) |
| High | 2,044 | (35) | 1,636 | (43) | 408 | (20) |
| Medium | 991 | (17) | 693 | (18) | 298 | (15) |
| Low | 1,399 | (24) | 789 | (21) | 610 | (31) |
| Very low | 659 | (11) | 311 | (8) | 348 | (17) |
| Trace | 681 | (12) | 346 | (9) | 335 | (17) |
| MTB NOT Detected | 65,554 | (92) | 8,389 | (69) | 57,165 | (97) |

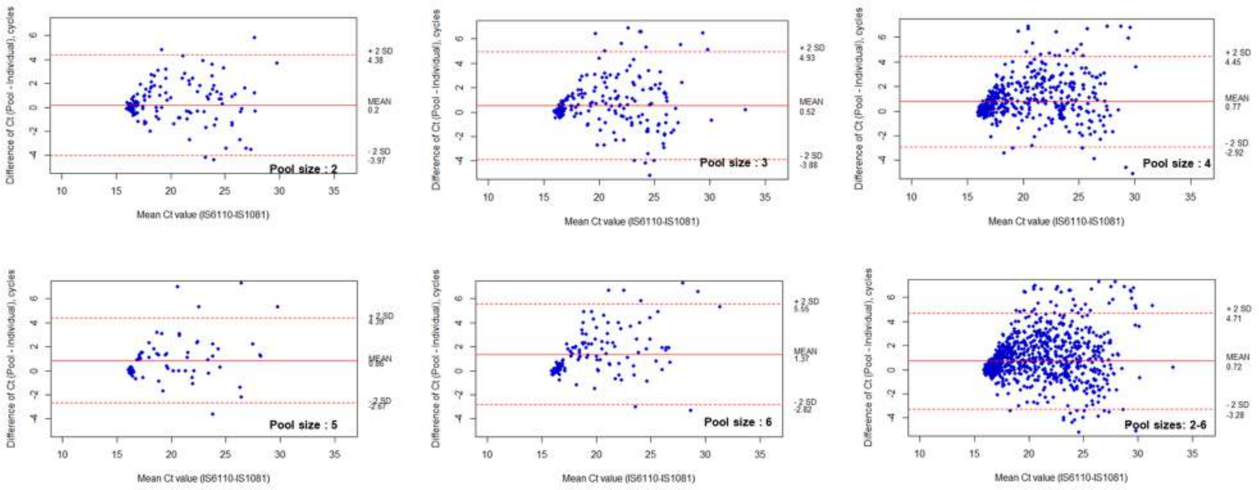

**Figure S2. Bland-Altman plots to compare cycle threshold values of specimens with positive results tested both in pools of size 2 and 6 and individually on the Xpert MTB/RIF Ultra assay**

The solid red line indicates the mean bias, and the dashed red lines indicate the 95% limits of agreement (LOA). Only specimens in pools with a single positive result are included. The average standard deviation of the bias across pools of size 2 to 6 is 2.0 cycles; the 11 pools of size 7 and 8 are not included due to the small sample size.

**Table S3. Summary of Bland-Altman agreement by pool size and overall**

| Pool size | Number of specimens | Mean bias (cycles) | 95% limits of agreement | Standard deviation of the bias (cycles) |
|---|---|---|---|---|
| 2 | 157 | +0.20 | +4.38 to -3.97 | 2.1 |
| 3 | 225 | +0.52 | +4.93 to -3.88 | 2.2 |
| 4 | 544 | +0.77 | +4.45 to -2.92 | 1.8 |
| 5 | 76 | +0.86 | +4.39 to -2.67 | 1.8 |
| 6 | 131 | +1.37 | +5.55 to -2.82 | 2.1 |
| Overall (2-6) | 1,133 | +0.72 | +4.71 to -3.28 | 2.0 |

The number of specimens included for each pool size, the mean bias, the 95% limits of agreement and the standard deviation of the bias from the Bland-Altman agreement analysis in Figure S2 are summarized in Table S3.
