## Supplementary material for "Implementation of pooled testing of sputum specimens from multiple individuals for rapid molecular detection of tuberculosis in Cameroon: retrospective evaluation of efficiency, cost, and instrument time to result": Job Aid

### Job Aid- Pooled testing of sputum samples for TB detection by the Ultra assay

**Scope:** Pooled testing can help reduce the time and cartridges required for diagnostic testing using the Xpert MTB/RIF Ultra assay. It is recommended that pooled testing be used for individuals at lower risk for TB. Pool testing should typically only be used in populations with a TB positivity rate of <25%.

**Note:** It is recommended that testing be performed with pools of either 2, 3, 4 samples or up to 8 samples as needed; If the positivity in the population is >10%, it may be preferable to use a pool size of 2; for positivity of <10%, typically a pool size of 3 to 6 preferred, depending on the lab positivity rate and individual patient characteristics, as well as test cartridge and module availability. Please contact the reference lab if you need guidance on how to choose the best pool size.

**Schematic overview of pooled testing:**

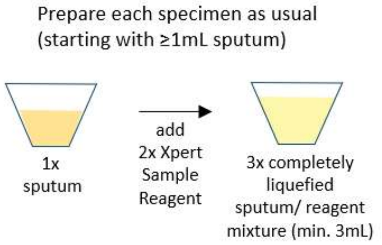


| Pool size | Volume of each sample/ reagent mixture to add to pool | Volume of pooled mixture for testing | Volume of individual sample/reagent mixture remaining (for individual re-testing if needed) |
|---|---|---|---|
| 2-4 | 1 mL | 2-4 mL | ≥2mL |
| >4 | 0.5 mL | >2.5 mL | ≥2.5mL |

Note: Person only submits one sputum specimen- typically no need for repeat collection

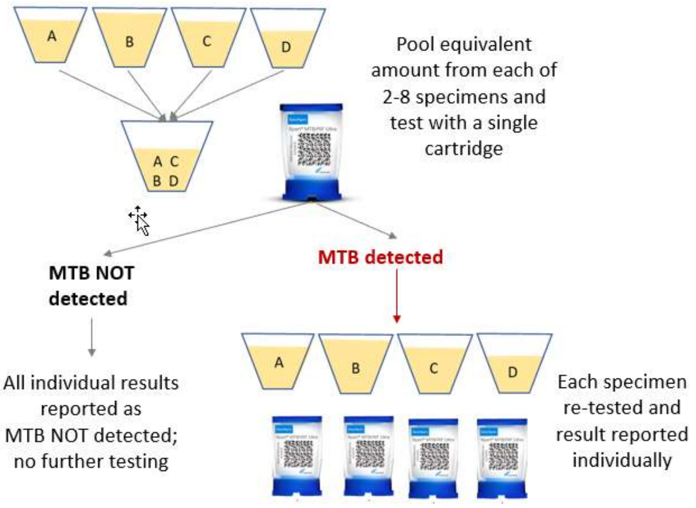


**Type of samples to be used for pooled testing and criteria for pooling**

- Only specimens sent for initial TB diagnosis
- Sputum specimens only
- Sputum specimens with a volume of 1.0 mL or more

**Type of specimens to be tested individually**

- Extrapulmonary or non-sputum pulmonary specimens
- Samples with a volume of less than 1.0 mL (because addition of 2x SR will not enable re-testing if pool is positive; if required, additional Xpert SR can be added to create a starting sample-reagent-mixture of 3mL in volume)
- Specimens that have an initial positive MTB or positive smear result and are sent for rifampicin resistance testing should be tested individually

**Materials needed:**

- Ultra cartridges (Xpert MTB/RIF Ultra cartridges)
- Xpert testing register (provided by reference lab).
- transfer pipettes (or repeat pipette with pipette tips); either provided with Ultra cartridges or provided separately.

- Xpert Sample Reagent (Xpert SR, provided with cartridges); may be used for more than one specimen if aseptic procedures are respected to avoid contaminating the reagent container between specimens by using a new transfer pipette for each specimen
- Timer (to time the duration of Xpert Sample Reagent in sputum sample)
- Sputum mugs (to create pooled specimen/reagent mixture).
- Bold marker (for writing testing numbers on pooled sputum mug and Xpert cartridges)
- GeneXpert instrument
- Waste disposal containers (for specimen preparation area and for Xpert testing area)
- Materials to disinfect working surfaces

**Procedure**

**IMPORTANT Note: Before starting testing, ensure your workspace is clean, free of clutter and ONLY specimens to be involved in the pooling are present in the working area. This is critical to avoid mistaking one sample for another and to avoid cross contamination.**

1. Collect/receive samples following routine protocols.
2. At check-in, sort request forms for those eligible for pooling.
3. Visually assess collected samples to identify those with sputum volume ≥1.0mL.

   **Notes:**

   a. If sputum volume is less than approximately 1.0mL, exclude this sample from the pool and process individually (this is to ensure there is enough specimen/reagent mixture left for subsequent individual testing as needed, if the pool result is positive or invalid/error/no result; alternatively, if required, add additional Xpert SR to bring the total volume up to 3mL).

   b. If the sample volume is ≥4mL, aliquot about 2mL into a new labeled sputum mug and use for testing to ensure sufficient Xpert samplereagent is available.

4. Create patient and specimen identification numbers and enter these into the Xpert register.

   **a.** For pooled testing, the Patient ID field (in both the Xpert register and the GeneXpert instrument) is filled using the first name of each patient in the pool separated by a period. e.g., for Mbacham Einstein, Ebot R, Tsala M, Penn M, this would be:

   Patient ID field is: Mbacham,Ebot,Tsala,Penn

   **Note:** For pools of six (6) use the initials of the patient e.g. for the example above the patient ID field will be ME,ER,TM,PM… if it were a pool of six.

   b. The Sample ID field is composed of the pooled ID (in both the GeneXpert instrument and Xpert register). This ID is created using "P(SN)" to indicate "pool serial number" followed by the last number of each specimen's record number, with a dash in between, e.g., for specimens 1BRH-00107, 1BRH-00129, 1BRH-00117, 1BRH-00189, this would be:

   **Sample ID: P001-7-9-7-9 (assuming this is pool number 1)**

   **Note:** Typically, the pool serial number should be assigned either from the beginning of when pooling started or at the beginning of the year and restart at the beginning of the next year. This pool serial number helps the laboratory to track the number of pools performed.

c. Enter the complete sample record number and initials of each patient’s name i.e 1BRH-00107-ME, 1BRH-00129-ER,1BRH-00117-TM, 1BRH-00189-PM into the Specimen ID column in the Xpert register

d. Enter each individual patient's name and sample ID tested as part of this pool in the next rows below the information for the pool test. The Xpert register for the above example should look like this:

| SN | Registration no. | Patient ID | Specimen ID (Sample ID) |
|---|---|---|---|
| 1 | - | Mbacham,Ebot,Tsala,Penn | P001-7-9-7-9 |
| 2 | 00107 | Mbacham Einstein | 1BRH-00107-ME |
| 3 | 00129 | Ebot R | 1BRH-00129-ER |
| 4 | 00117 | Tsala M | 1BRH-00117-TM |
| 5 | 00189 | Penn M | 1BRH-00189-PM |

5. Label a new sputum mug with the Pool ID, for example, P001-7-9-7-9. This container will be used to create the pooled sample/reagent mixture. Place this labeled container behind the specimens to be pooled.

6. Prepare each of the individual samples to be pooled according to the normal specimen preparation procedure for the Xpert MTB/RIF Ultra assay, i.e.

   i. For each specimen: using a new transfer pipette (or pipette with tips), add 2 volumes of Xpert Sample Reagent to 1 volume of sample and then close the container. For example, if the sputum volume is 1.5 mL, add 3.0 mL for a total volume of 4.5 mL (note that during pooling, you will add 1.0 mL to the specimen pool, so that 3.5 mL will remain in case the pool Ultra result is positive and you need to re-test that sample individually for Ultra). (Note: If the sputum specimen volume is close to 1mL, add slightly more than 2mL Sample Reagent. This is to ensure you will have slightly more than 2mL of the sample/reagent mixture remaining in case you need to re-test individually).

   **Important Notes:**

   - Be cautious not to contaminate the remaining Xpert Sample Reagent and close it immediately after use. If the Sample Reagent is only pipetted with a dedicated pipette and never contacts the specimen, specimen container or a contaminated pipette, it can be used as a source of Sample Reagent for other specimens. This will help to extend the availability of Sample Reagent for specimen pooling.
   - If using a transfer pipette, use a new pipette for each specimen and keep the pipette in the pipette plastic between steps. Make sure you put back the used transfer pipette in the pipette plastic and keep it beside the sample as it will also be used to transfer the sample/reagent mixture to the pool container for that specimen.
   - If using a standard pipette, discard the used pipette tip immediately after use.

   ii. Mix the specimen and Xpert Sample Reagent thoroughly by shaking vigorously 10 to 20 times. Incubate for 10 minutes at room temperature.

   iii. Mix thoroughly and incubate for an additional 5 minutes at room temperature.

   iv. Visually inspect the specimen/reagent mixture. If it is completely liquefied, proceed to the next step. If it is only partially liquefied, mix again thoroughly and let stand for an additional 5 minutes or until completely liquefied. Avoid pipetting solids or debris into the Ultra cartridge to avoid errors.

7. Once each of the individual sample mixtures are liquefied, create the pooled sample by adding each sample/reagent mixture to the labeled sputum mug dedicated to the pooled sample. Carefully open the individual sputum mug and, depending on the pool size, transfer an amount from each sample as indicated on the table below into the dedicated pooled sputum mug using the same transfer pipette used in step 6(i) above. Close the lid of the individual sputum mug before opening the next sputum mug for the next sample.

| Pool size | Volume of each sample/reagent mixture to add to pool | Volume of pooled mixture for testing | Volume of individual sample/reagent mixture remaining (for individual re-testing if needed) |
|---|---|---|---|
| 2-4 | 1 mL | 2-4 mL | ≥2 mL |
| 5-8 | 0.5 mL | >2.5 mL | ≥2.5 mL |

8. Repeat step 7 for each sample/reagent mixture to be included in the pool.

9. Set aside the sputum mug containing the individual samples/reagent mixtures for later use if necessary (if the pooled test result is positive or invalid, each sample must be retested individually).

10. Label the cartridge to be used for pool testing with the pool information from the sample ID field (e.g., P001-7-9-7-9).

11. Using one of the transfer pipettes used for the individual samples, aspirate the liquefied sample pool just above the line on the pipette (just above 2 mL). If there is insufficient sample volume, do not proceed with testing. (Note: If the test is run with <=2mL, it may generate an error.)

12. Perform an Xpert Ultra test using the pooled sample according to the Cepheid protocol. Verify that the cartridge is labeled with the same Pool ID as the one on the pooled sample container (For example, P001-7-9-7-9).

13. To enter specimen information into the GeneXpert instrument, use the following: For the "**Notes**" field in the GeneXpert instrument, using the Barcode scanner of the GeneXpert instrument, point it on the QR code of the first participant in the pool to scan the QR code. Repeat this for all the participant QR codes in the pool. Put a dash (-) and add the initials of the participants corresponding to the scanned QR code as shown below. Note; In case a QR code is not used, enter the laboratory unique ID in the notes field instead; this is important to enable linkage to a subsequent individual test if needed.

| Field type | Lab Register | GeneXpert instrument |
|---|---|---|
| Patient ID | Names (e.g. Mbacham,Ebot,Tsala,Penn) | Names (e.g. Mbacham,Ebot,Tsala,Penn) |
| Sample ID | P001-7-9-7-9 | P001-7-9-7-9 |
| Notes | N/A (see 4d above) | 1BRH-00107-ME<br>1BRH-00129-ER<br>1BRH-00117-TM<br>1BRH-00189-PM |

14. When the results are ready, save them using the steps below:

**a. For a positive pool (MTB Detected)**

i. If the pooled sample result is "MTB Detected," with any grade, record the result in the Xpert register with the pooled sample ID as MTB Detected and retest all four samples individually.

ii. Repeat the test for each patient in a positive pool individually, for each sample:

- Record the serial number in the Xpert register for each line where an Ultra cartridge is used.
- Label a new cartridge with the individual sample ID, e.g., 1BRH-00107-ME, as on the individual sputum mug.
- Using a new transfer pipette, aspirate the liquefied individual specimen/reagent mixture reserved in step 7 just above 2 mL and transfer to a new Ultra cartridge. Label the cartridge with the patient registration number and initials.
- Enter the patient information into the GeneXpert instrument (as appropriate), using the following:

| | |
|---|---|
| Patient ID field | Patient name, e.g. Mbacham Einstein |
| Sample ID field | 1BRH-00107-ME |

iii. Once the individual test results are available, record the individual results in the Xpert register with the individual sample ID as obtained from the instrument. Note: Each time an Ultra cartridge is used, the serial number column is filled in. This allows the laboratory to track the use of the Ultra cartridges.

iv. After repeating the individual tests, complete the Xpert results log with the individual test results as shown below.

| SN | Sample ID | Xpert Ultra results | If MTB (RR-IND, NOT) |
|---|---|---|---|
| 1 | P001-7-9-7-9 | MTB Detected Trace | IND |
| 2 | 1BRH-00107-ME | MTB Not Detected | N/A |
| 3 | 1BRH-00129-ER | MTB Very Low | NOT |
| 4 | 1BRH-00117-TM | MTB Not Detected | N/A |
| 5 | 1BRH-00189-PM | MTB Not Detected | N/A |

v. Prepare individual results to be given to the client when they come to pick up their results.

**Note:** All samples in a positive pool should be tested to identify the positive before results from that pool are given out.

**b. For a pool with INVALID result or ERROR, follow the same steps as for a positive pool.**

If the `pooled sample gives an error or invalid result, record this result in the Xpert register next to the pooled sample ID as "Error", or "Invalid", or "No Result", depending on the result given by the instrument and repeat the test individually following the same steps as for a positive pool. **Note:** You may decide to repeat a pool with an error or invalid result if you consider that after performing a pool repeat you will still have enough samples to perform individual tests for each specimen in the case of a positive.

c. **For a negative pool (MTB Not Detected) follow the steps below.**

i. If the test result is "MTB Not Detected", all three samples are MTB not detected. Record the result in the logbook with the pooled sample ID and the individual sample IDs as MTB not detected.

Example of a record for a negative pooled test result (Note: The serial number column is only filled when a cartridge is used. This helps the laboratory keep track of Ultra cartridge usage.)

| SN | Sample ID | Xpert Ultra results | If MTB (RR-IND, NOT) |
|---|---|---|---|
| 1 | P001-7-9-7-9 | NOT | N/A |
| | 1BRH-00107-ME | NOT | N/A |
| | 1BRH-00129-ER | NOT | N/A |
| | 1BRH-00117-TM | NOT | N/A |
| | 1BRH-00189-PM | NOT | N/A |

ii. Prepare the individual results for the patient and give them to the patient when they come to pick up their results.

**Notes:**

- The purpose of individually retesting each specimen from a positive pool is to identify which specimen(s) is/are positive; If the individual specimen result identifies one or more positive specimens, report the individual specimen result obtained by following the normal procedure for reporting a positive result.
- Any positive result (MTB Detected- Trace, Very Low, Low, Medium, High) indicates that the patient should be started on TB treatment immediately according to national guidelines.
- While it is rare, there may be times when you test each sample from a positive pool individually and none of them have a result of "MTB Detected." If all individual samples have a result of "MTB NOT detected," collect a second sample from each patient and retest each individually. If the test result identifies the sample with "MTB Detected," report the result as usual. If retesting of all the specimens in the positive pool does not result in any "MTB detected" result, then report each result as "MTB NOT detected." Add each of these patients to the patient follow-up record and continue to follow each patient until they are better, including providing additional Ultra tests as needed.
- It is very important to test the individual specimens from any positive pool within 4 hours of adding the sample reagent. If this cannot be done, store the specimen/reagent mixture in the refrigerator and test within 24 hours. Failure to test within the stated time may lead to degradation of the DNA and could result in a false positive pool, for example.
- Contact the reference laboratory if you have any questions or require further information.
